# How can telehealth be used to enhance the quality of care in home-based pediatric palliative care: protocol for a systematic literature review

**DOI:** 10.1101/2025.10.08.25337424

**Authors:** Christopher Adlung, Christa Niehot, Carine van Capelle

## Abstract

Pediatric palliative care (PPC) is a specialized medical approach aimed at improving quality of life of children with life-limiting and life-threatening conditions. It addresses the needs of both the child and family, and preferably starts immediately after a palliative diagnosis with access to specialized care, effective symptom management, and psychosocial, spiritual and emotional support. Unfortunately, receiving PPC is often challenging due to the varied and complex nature of conditions and the difficulties in reaching all patients who require care. Telehealth offers a promising solution by enabling virtual access to interdisciplinary teams, facilitating real-time consultations, extending care into the home, educating professionals across regions, and fostering consistent, collaborative, patient- and family-centered PPC. As this requires a seamless integration into the daily routines of all relevant stakeholders, telehealth may raise complexity in terms of privacy, data protection and regulatory compliance. Nevertheless, studies indicate that parents and patients are open to using telehealth applications. Evidence on the quality of care provided through digital health interventions in PPC remains limited. Therefore, the aim of this study is to investigate how telehealth can enhance the quality of care in home-based PPC settings, not only for patients, but also for families and healthcare professionals. Findings from our review will contribute to a deeper understanding of how patients and families can receive timely and equitable palliative care regardless of their location, ultimately informing future models of digital health interventions in PPC.

**Methods and analysis:** We will follow the Preferred Reporting Items for Systematic Reviews and Meta-Analyses (PRISMA) guideline and perform a systematic search across Medline ALL, Embase, Web of Science Core Collection, Cochrane Central Register of Controlled Trials, and Google Scholar. Results will be displayed by the PRISMA flow diagram, visually summarizing the screening process. We will consider peer-reviewed studies without language, time or geographic restrictions and manage extracted data using Covidence. The search will be conducted by an information specialist and data synthesis will be documented via a data extraction table. Two reviewers will screen and extract data independently, with a third reviewer resolving discrepancies. We will present a narrative synthesis, using clear language, defining key terms, and following open-access standards to ensure accessibility for non-expert audiences.

**Ethics and dissemination:** No primary or clinical data will be collected. The review findings will be shared via peer-reviewed journal publications, conferences, and stakeholder meetings, and video presentations.

**PROSPERO registration number:** CRD420251035350

**STRENGTHS AND LIMITATIONS OF THIS STUDY:**

- We will identify and highlight important new developments that may enhance the current understanding of the potential role of telehealth interventions in PPC.
- We will explore how quality of care and respective metrics are described in the context of home-based PPC.
- We will investigate the functionalities and features deployed in telehealth solutions.
- We will identify barriers and facilitators to adopt a home-based telehealth model for PPC.

## INTRODUCTION

### Promise of telehealth to transform PPC

According to the World Health Organization, pediatric palliative care (PPC) is “the approach to prevent and relief suffering of pediatric patients and their families facing the problems associated with life-threatening illness. These problems include the physical, psychosocial, social and spiritual suffering of patients and their family members”.^1^ The exact number of children in need of PPC is currently unknown. Estimates vary considerably across countries, with the highest rates reported in low-resource settings. Connor et al. assessed that more than eight million children worldwide are in need of specialized PPC.^2^

The spectrum of care required can be categorized based on the child’s diagnosis and disease progression^3^, developmental stage, psychosocial, spiritual and cultural needs^4^, health literacy, i.e. interaction between the individual and the health care system^5^, and access to adequate resources.

The care journey of children with life-limiting diseases begins with early identification of PPC needs and timely integration of palliative care into their general pediatric care. As the child’s condition evolves, early referral to specialized palliative care teams is crucial when complex needs emerge.^6^ PPC teams are multidisciplinary groups that provide comprehensive, specialized care, coordinate services, and offer support centered on the needs of both patients and their families. The continuity of care they provide, preferentially delivered in a home-based environment, is of utmost importance. As the family of seriously ill children face greater psychological distress, poorer mental health, and worse physical well-being compared to their peers,^7,8^ the need to rely on information, advice and support from specialized pediatric care clinicians is even more important.^9^ In practice, PPC requires a delicate balancing between clinical decision-making and the values, preferences, and best interests of the child and their family.

Unfortunately, the clinical care journey is still characterized by diverse challenges, including patient and population diversity, prognostic uncertainty, the need for integration of PPC into the broader healthcare system, a preference for home-based care, difficult clinical decision-making, logistical challenges^10^, limited pediatric palliative resources alongside significant geographic distances.

Considering these challenges, it is essential to explore how the PPC journey can be enriched to better meet the needs of patients and families. Telehealth has the potential to enhance communication and shared decision-making between healthcare professionals and patients, while improving access to care. This can be achieved through virtual collaboration, leveraging interdisciplinary expertise, and offering real-time consultations within the home environment, all of which support consistent, collaborative, and family-centered PPC. Other telehealth modalities, like telemonitoring, have the ability to collect and share health information, not only enhancing communication between health care professionals and patients, but also improving symptom management and engagement of patients.

Telehealth, being embodied within the WHO Global strategy on digital health 2020-2025, relates to “*the field of knowledge and practice associated with the development and use of digital technologies to improve health*”, should be “*developed with principles of transparency, accessibility, scalability, replicability, interoperability, privacy, security and confidentiality.*”^11^ Currently, information is mostly delivered to patients and families by PPC teams in hospital settings and home-based environments in an analogue and face-to-face format. The promise of telehealth is to enable, to control and to enrich clinical encounters with the direct transfer of information via synchronous and asynchronous ways.

This possesses the potential to provide and access care, support and continuity of care^12^, without the need for families and patients to travel to the hospital, search for parking, and endure long waiting times. Reflecting on the holistic approach of PPC, studies also demonstrate the ability of digital technologies to deliver support, extending the medical care of these patients, while including emotional support for patients and families.^13^ Telehealth and telemonitoring offer seamless integration into families’ daily routines, with simple, user-friendly tools. By minimizing complexity, telehealth technologies can enable easy access to care, which could be of extra value in the context of immediate deterioration, fostering greater engagement and continuous communication with care teams.

One significant gap which telehealth could fill is the restricted availability of specialized PPC teams. Families often have to turn to local pediatric services, which may not be adequately equipped to meet the specialized patient’s needs. Telehealth addresses this gap by facilitating access to expertise in real-time, allowing families and local care teams to connect with PPC professionals regardless of geographical barriers.^13^

### Limitations of telehealth in PPC

Telehealth solutions are confronted with various challenges, including the perception of intrusions in home environments and reluctance among healthcare professionals^14^, accessibility barriers in the enrollment of telehealth research^15^, technical difficulties and connectivity issues^16^, socio-economic differences, and limitations in capturing the subjective reality of the patient in clinical encounter.^17^

Participants of telehealth services expressed little concern over privacy, comfortably sharing sensitive data digitally, unlike the privacy anxieties often seen in adult palliative care settings with telehealth tools.^18^ However, there is concern that telehealth may weaken essential human connections between healthcare professionals and families.^19,20^ This concern aligns with similar research findings, suggesting that telehealth, despite its benefits, cannot replicate the comprehensive depth and quality of face-to-face care.^12^

Despite the nature of telehealth, and its potential benefits for families in PPC, literature describing its use in the pediatric population and the impact on quality of care remains limited. Studies on home-based telehealth were reviewed, examining different outcome measures such as quality of life, anxiety, barriers, feasibility and acceptability.^9,13^ Unfortunately, these outcomes have often been constrained by various methodological and practical challenges, and did not describe the direct impact on quality of care.

### Importance of quality of care and outcome measures in PPC

As outlined by Mulder et al., quality of care is defined as “whether individuals can access the health care structures and processes of care which they need and whether the care received is effective”.^21,22^ The goal of assessing quality of care is to improve child and family outcomes, such as quality of life and care experiences.^23^ When implementing new interventions, quality of care measurement is even more important to secure effectiveness, safety and acceptability while comparing the new intervention with traditional care. Quality of care measurement relies on Quality Indicators (QI). Health care QIs are a type of performance measure borrowed from the industrial process control systems literature. They are designed to compare actual patient care to ideal criteria.^24^

While the specialized field of PPC is actively working towards the development of standardized quality metrics, a consensus remains pending.^25,26,27^ Some researchers initiated to explore the landscape of PPC quality metrics:

Hommes et al. identified quality measures recognized by key stakeholders in PPC, both in hospital and in home-based care. Their work resulted in a measurement framework of seven domains, (i) alleviation of distressing symptoms, (ii) structures and processes of care, (iii) health care utilization, (iv) location of death and bereavement care, (v) patient and family experiences, (vi) psychological and spiritual care, and (vii) cultural, ethical, and legal considerations.^28^

According to Boyden et al., high-quality measures in PPC, should be population-based and account for the heterogeneous needs of patients and families. They should include clearly defined criteria for interpretation and benchmarks for assessing care quality.^23^ A recent scoping review from Ruiz-Gil and Ródenas-Rigla outlined three major areas, (i) structure and process of care, (ii) psychological aspects of care, and (iii) care of patients approaching the end of life.^29^ Alongside these metrics, the study emphasizes the need for institutional-level changes to healthcare infrastructure, aiming to improve both the quality of care and the effectiveness of services provided.

The primary objective of this systematic review is to give an updated and comprehensive overview of current knowledge on the potential role of telehealth interventions in PPC. We seek to identify and highlight important new developments that may enhance the current understanding in this field of expertise. A second aim is to give an insight into qualitative and quantitative metrics currently used to assess quality of care, ideally identifying quality indicators that can actively contribute to future implementation and adoption of telehealth solutions in PPC.

## Foundational concepts and definitions

### PEDIATRIC PALLIATIVE CARE

According to the WHO, “palliative care for children is the active total care of the child’s body, mind, and spirit, and also involves giving support to the family. Effective palliative care requires a broad multidisciplinary approach that includes the family and makes use of available community resources, and it can be successfully implemented even if resources are limited”.^30^

### TELEHEALTH

In addition, the WHO defines telehealth as the “delivery of health care services, where patients and providers are separated by distance”.^31^ In PPC, enhancing the quality of care has been addressed in form of improved access to care^32^, communication^15^, exchange of information^33^, technological trust^12^, shifts in power dynamics in favor of equal and democratizing healthcare encounters^34^, and quality indicators^35^.

### QUALITY OF CARE

Analyzing PPC and telehealth through the lens of quality of care reveals a lack of consensus on its definition and the establishment of metrics. To ensure comprehensive description of articles addressing quality of care, we will incorporate a broad interpretation of the concept, encompassing the following care domains, (i) structure (physical and staff characteristics), (ii) process (clinical and inter-personal care), and (iii) outcome (health status and user evaluation).

## Objectives

In this systematic review, we aim to synthesize and critically evaluate the growing body of literature based on the following research questions:

RQ1: To what extent does the use of telehealth interventions impact the quality of care in home-based paediatric palliative care settings?

RQ2: Which medical use cases are addressed within the care pathways?

RQ3: Under what circumstances is telehealth being used in paediatric palliative care at home?

RQ4: Which key features and functionalities exist in home-based telehealth interventions?

RQ5: What are the barriers and facilitators to use telehealth interventions in paediatric palliative home care?

RQ6: What conceptual frameworks exist for adopting telehealth into paediatric palliative care at home?

RQ7: What are the metrics for evaluating telehealth interventions in PPC?

## METHODS AND ANALYSIS

The methods in this review are reported based on the Preferred Reporting Items for Systematic Reviews and Meta-Analyses (PRISMA) Checklist^36^ and the Prisma-S extension to the PRISMA Statement for Reporting Literature Searches in Systematic Reviews^37^. An exhaustive search strategy was developed by an information specialist (CN) in cooperation with the lead author (CA). The search was developed in Embase.com, optimized for sensitivity and then translated to other databases following the method as described by Bramer et al^38^. The search was carried out in the databases Medline ALL via Ovid (1946 to Daily Update), Embase.com (1971-present), Web of Science Core Collection (Science Citation Index Expanded (1975-present); Social Sciences Citation Index (1975-present); Arts & Humanities Citation Index (1975-present); Conference Proceedings Citation Index-Science (1990-present); Conference Proceedings Citation Index-Social Science & Humanities (1990-present) and Emerging Sources Citation Index (2015-present) and the Cochrane Central Register of Controlled Trials via Wiley (1992-present). Additionally a search was performed in Google Scholar from which the 100 highest-ranked references were downloaded using the software Publish or Perish^39^, after the original search was performed in April 2025.

The search strategies for Medline and Embase used relevant thesaurus terms from Medical Subject Headings (MeSH) and Emtree respectively. In all databases, terms were searched in titles, abstracts and author keywords. The search contained terms for (i) telemedicine, (ii) palliative care and (iii) children. Terms were combined with Boolean operators AND and OR and proximity operators were used to combine terms into phrases. The full search strategies of all databases are available in the appendix section. No study registries were searched, but Cochrane CENTRAL retrieves the contents of ClinicalTrials.gov and World Health Organization’s International Clinical Trials Registry Platform. The reference lists of retrieved non-included relevant review articles and of the included references, as well as articles citing these papers have been scanned for relevant references missed by the search. No authors or subject experts were contacted and no extra unindexed journals in the field were browsed. The references were imported into EndNote and duplicates were removed by an information specialist (CN) using the method as described by Bramer et al.^38^ Two reviewers (CC and CA) will independently first screen titles and abstracts in Covidence and in the second stage full text. Any discrepancies in the verdict, will be resolved by discussion with a third reviewer. The paper was registered with the International Prospective Register of Systematic Reviews (PROSPERO) on August 7^th^, 2025 under the registration number CRD420251035350.

### Exclusion criteria

The inclusion and exclusion criteria will be guided by the PIO framework.^40^ Findings will be synthesized based on the following:

- Population (P): Pediatric patients and families receiving palliative care in a home-based environment.
- Intervention (I): Telehealth.
- Outcome (O): Enhanced quality of care in home-based environment.

Table 1 outlines the exclusion criteria.

**Table 1.** Exclusion criteria

| Priority | Exclusion Criteria | Description | Example |
| --- | --- | --- | --- |
| 1 | Non-eligible document types | • Review<br>• Policy documents and guidelines, e.g. WHO framework concepts<br>• Commentary<br>• Editorial working papers<br>• Posters<br>• Pre-prints: Documents that have not been reviewed yet by a scientific audience.<br>• Conference papers, where only abstract is available.<br>• Letters<br>• Written forms of communications among editors<br>• Entire books | n.a. |
| 1 | Unavailable full text | If no full text is available, documents will be excluded from review. | n.a. |
| 2 | Subject incompatibility for PPC | If none of the definition criteria for PPC is met, the study will be excluded. If partial criteria for PPC are met, the article will be included. | Study solely focus on adult population. |
| 2 | Telehealth | Should the patient receive treatments in a hospital setting, articles will be excluded. | Hospital-based PPC care. |

### Inclusion criteria

Table 2 outlines the inclusion criteria.

**Table 2.** Inclusion criteria

Inclusion criteria Table 2 outlines the inclusion criteria.
| Table 2 Inclusion criteria |  |  |  |
| --- | --- | --- | --- |
| Priority | Inclusion Criteria | Description | Example |
| 1 | Document type | • Empirical studies<br>• Unindexed material<br>• Quality control studies<br>• Single case studies<br>• Multiple-site studies about telehealth according to the WHO definition<br>• Book chapters | n.a. |
| 1 | Languages | • Inclusion of all languages | n.a. |
| 2 | Telehealth | • Any concept related to the notion of telehealth.<br>• Typically, telehealth interventions cover the notion of geographical separation between patient and provider.<br>• Articles will be included if telehealth interventions include the notion of medical or educational services.<br>• Articles will be included in case telehealth intervention is performed in home-care setting. | e-health, telemedicine, e-health, m-health, and u-health, et cetera. |
| 2 | Adoption and implementation factors | Article will be included if they address the notion of adoption, implementation, initiation, diffusion, or scale-up. | Adoption of telehealth intervention. |
| 2 | Patient population | We will include articles in which patients are between 0-18 years old. | Patients between the age of 0-18 years old. |
| 2 | Quality of care metrics | Articles will be included if the definition of quality of care is met. | Process or structural measures of quality of care. |

### Information sources and search strategy

To answer the RQs, our search strategy encompasses key concepts through a series of Boolean operators (“AND”, “OR”). The search strategy was developed in collaboration with an information scientist, and we will search the following databases: Medline ALL, Embase, Web of Science Core Collection, Cochrane Central Register of Controlled Trials, and Google Scholar. The full list of search terms can be found in the appendix section.

Table 3 outlines the expertise and roles of all authors.

**Table 3.** Expertise and roles of authors.

| Author | Role | Expertise |
| --- | --- | --- |
| CA | Lead author and guarantor of review. | PhD researcher focusing on implementation, adoption, and scale-up of digital health interventions, in specific telemedicine. |
| CN | Co-author, search string compilation and database extraction. | Information specialist, specialized in composing search strings used in various medical databases. |
| CC | Co-author, domain expert in PPC. | Pediatrician specializing in metabolic, genetic, and palliative care. |

### Screening and data collection process

All decisions regarding study inclusion in the review will be based on the eligibility (inclusion and exclusion) criteria. We will manage all screening records, full-text review, risk of bias assessment, and conflict resolution using Covidence. The study selection process will involve the following stages: (i) initial screening of titles and abstracts, (ii) full-text review for eligibility, and (iii) final inclusion decision. At each stage, two reviewers will independently screen records and assess eligibility, disagreements will be resolved by consensus. In line with the study selection process, data will be extracted from all records eligible for final inclusion by two independent reviewers. Reviewer disagreements will be resolved through discussion, with consensus documented in the final data extraction table. Two authors will screen 10% of articles for consistency, then proceed to screen all remaining articles. Full-text screening will be conducted by reviewers, with bi-weekly meetings and consensus meetings.

### Data items and review outcomes

Given the outlined research questions, we will aim to extract the following data items. Table 4 outlines the data items to extract.

**Table 4.**
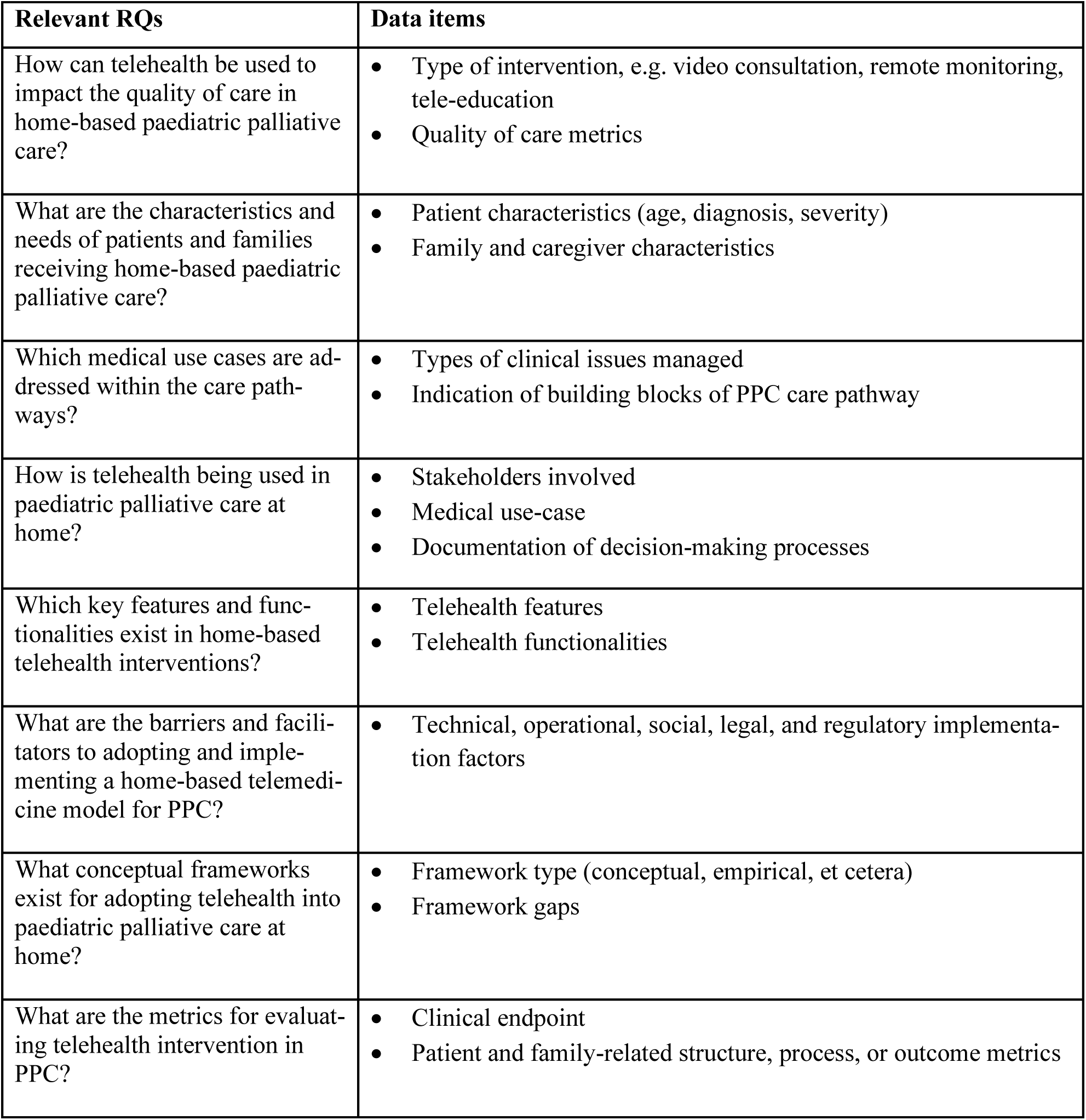
Data items to extract

| Relevant RQs | Data items |
| --- | --- |
| How can telehealth be used to impact the quality of care in home-based paediatric palliative care? | • Type of intervention, e.g. video consultation, remote monitoring, tele-education<br>• Quality of care metrics |
| What are the characteristics and needs of patients and families receiving home-based paediatric palliative care? | • Patient characteristics (age, diagnosis, severity)<br>• Family and caregiver characteristics |
| Which medical use cases are addressed within the care pathways? | • Types of clinical issues managed<br>• Indication of building blocks of PPC care pathway |
| How is telehealth being used in paediatric palliative care at home? | • Stakeholders involved<br>• Medical use-case<br>• Documentation of decision-making processes |
| Which key features and functionalities exist in home-based telehealth interventions? | • Telehealth features<br>• Telehealth functionalities |
| What are the barriers and facilitators to adopting and implementing a home-based telemedicine model for PPC? | • Technical, operational, social, legal, and regulatory implementation factors |
| What conceptual frameworks exist for adopting telehealth into paediatric palliative care at home? | • Framework type (conceptual, empirical, et cetera)<br>• Framework gaps |
| What are the metrics for evaluating telehealth intervention in PPC? | • Clinical endpoint<br>• Patient and family-related structure, process, or outcome metrics |

### Risk of bias in studies

We will assess the methodological quality of the included studies using the Mixed-Method-Appraisal-Tool (MMAT).^41^ This tool is designed to assess the methodological quality of empirical studies, incorporating various study designs. The selection of the MMAT will be based on the study design of each included study. Quality assessments will be conducted independently by the reviewers. Discrepancies will be resolved via consultation.

### Synthesis methods and evidence assessment

The extracted data from identified articles will first be summarized and synthesized in an extraction table in MS Excel. This process involves organizing the information in a structured and systematic way, improving the summary of the data. The synthesized data from the extraction table will then form the basis of the final results section of the review. A narrative summary of the findings will be prepared, integrating the outcomes from all included studies. This narrative will be aligned with the RQs to ensure a clear connection between the synthesis and the overarching goals of the review. Additionally, the narrative synthesis will be supported by tables that present the extracted data from each study in a concise and accessible format. Underlying patterns could reveal quality of care metrics that are relevant for assessing potential enhancement of telehealth interventions in PPC. By analyzing these patterns, we may identify key indicators, reflecting the impact of telehealth on patient outcomes in home-based settings.

## DISCUSSION

In our review, we first aim to expand and update the current literature on telehealth interventions in PPC. Second, we aim to find criteria, including quality of care metrics, that are distinctive to the heterogenous pathway of children with PPC needs. By also adding results on adoption and implementation factors, i.e. technical, operational, social, legal, and regulatory, we are outlining the current state of telehealth in PPC, and we aim to extent reach with the final goal to inspire other healthcare providers. In specific, we anticipate that the results will guide healthcare providers in conceptualizing and developing telehealth solutions, promoting the quality of care of home-treated PPC patients. Ultimately, our work seeks to increase access for patients to receive the specialized care they are entitled to.

### Limitations

This systematic literature review will encompass qualitative and quantitative studies to ensure a comprehensive evidence base. Given the interdisciplinary nature of the topic, we anticipate a high level of heterogeneity. To capture evidence from all relevant fields, we will include studies in all languages and incorporate snowballing. Snowball sampling will be used in both forward and backward directions to ensure a comprehensive exploration of relevant sources. We will focus on a limited number of primary sources and prioritize key studies, while addressing the challenge of varying levels of analysis across sources.

### Author affiliations

- Department of Multi-Actor Systems, Faculty of Technology, Policy and Management, Delft University of Technology, Delft, Netherlands
- Literature Searches Support, Dordrecht, The Netherlands
- Sophia Children’s Hospital, Erasmus University Medical Center Rotterdam, The Netherlands

## Data Availability

All data produced in the present work are contained in the manuscript

## Acknowledgements

n.a.

## Collaborators

The following coauthors and collaborators are included on behalf of Delft University of Technology and Medical Library, Erasmus University Medical Center Rotterdam, The Netherlands.

## Contributors

CA: Lead author.

CN: Co-author, search string compilation and database extraction.

CC: Co-author, domain expert in pediatric palliative care, and guarantor of review.

### Funding

This research is supported by Convergence Healthy Start, a program of the Convergence Alliance - Delft University of Technology, Erasmus University Rotterdam and Erasmus Medical Center - to improve the future of new generations. Grant number HSF25_05.

### Patient and public involvement

No patients and/or the public were involved in the design, conduct, reporting, or dissemination plans of this research.

### Patient consent for publication

Not applicable.

### Provenance and peer review

Not commissioned; externally peer reviewed.

## Supplemental material

This content has been supplied by the authors. It has not been vetted by BMJ Publishing Group Limited (BMJ) and may not have been peer-reviewed. Any opinions or recommendations discussed are solely those of the authors.

## Appendix

20250416 Christopher Adlung

Telemedicine palliative child

SR

Trials: no

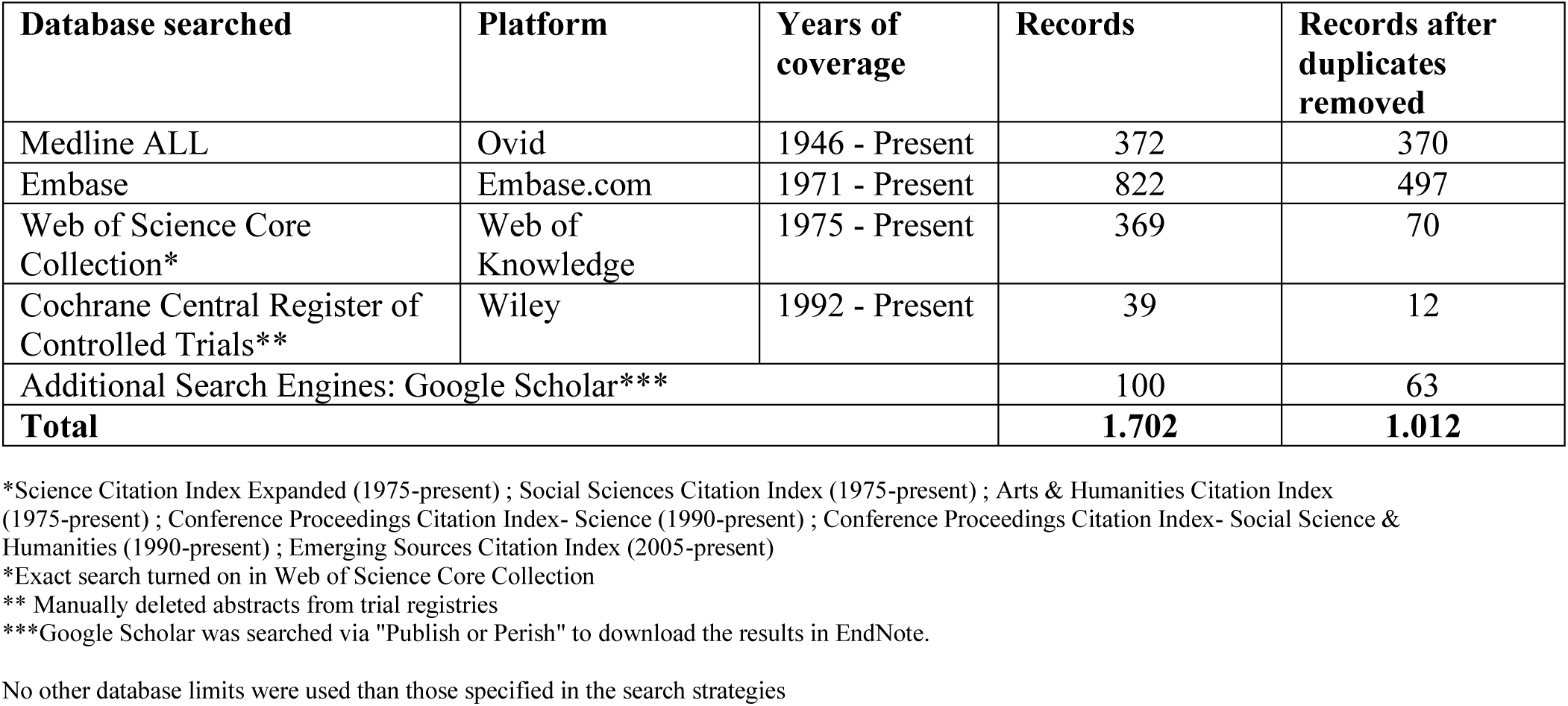

### Medline

(Telemedicine/ OR Remote Consultation/ OR *Telecommunications/ OR exp Digital Health/ OR exp Mobile Applications/ OR Telephone/ OR Cell Phone/ OR Videoconferencing/ OR Internet-Based Intervention/ OR Text Messaging/ OR (telehospital* OR tele-hospital* OR e-health* OR ehealth* OR (electronic-health* NOT electronic-health-record*) OR mhealth* OR m-health* OR telehealth* OR tele-health* OR telemedic* OR tele-medic* OR telemonitor* OR tele-monitor* OR telecare OR tele-care OR telephone* OR telepresence* OR tele-presence* OR tele-referral* OR telereferral* OR teleconsultat* OR ((digital* OR remot* OR electronic* OR virtual* OR mobile OR web-based OR webbased OR internet-based* OR internetbased*) ADJ3 (health* OR intervention* OR consult* OR diagno* OR medicine*)) OR health-information-technolog* OR health-IoT OR h-IoT OR uhealth OR ((remot*) ADJ3 (monitor*)) OR ((smart*) ADJ3 (health*)) OR ((health OR medical) ADJ2 (internet-of-things)) OR ((digital* OR remot* OR electronic* OR virtual* OR mobile) ADJ3 (distanc*) ADJ3 (practice*)) OR smartphone OR cellphone* OR ((smart* OR cell*) ADJ2 (phone*)) OR videoconferenc* OR video-conferenc* OR ((video*) ADJ3 (consult*)) OR ((mobile*) ADJ3 (app* OR health)) OR ((home*) ADJ3 (monitor*)) OR ((remote*) ADJ3 (care*)) OR tele-mental* OR telemental*).ab,ti,kf.) **AND** (exp Palliative Care/ OR exp Terminal Care/ OR (palliat* OR ((symptomatic*) ADJ3 (treat* OR therap*)) OR end-of-life* OR ((terminal*) ADJ3 (care*))).ab,ti,kf.) **AND** (exp Child/ OR exp Infant/ OR exp Pediatrics OR (pediatr* OR paediatr* OR neonat* OR neo-nat* OR child* OR toddler* OR boy OR boys OR girl OR girls OR infant OR infants OR baby OR babies OR newborn*).ab,ti,kf.)

### Embase

(’telemedicine’/de OR ’video consultation’/exp OR ’telediagnosis’/de OR ’teleconsultation’/de OR ’telecare’/exp OR ’telehealth’/exp OR ’telemonitoring’/exp OR ’telecommunication’/de/mj OR ’digital health’/exp OR ’digital health technology’/exp OR ’digital health intervention’/exp OR ’electronic consultation’/exp OR ’mhealth’/exp OR ’mobile health’/exp OR ’mobile health application’/exp OR ’mobile technology’/exp OR ’mobile application’/exp OR ’telephone’/de OR ’mobile phone’/exp OR ’videoconferencing’/de OR ’web-based intervention’/de OR ’text messaging’/de OR (telehospital* OR tele-hospital* OR e-health* OR ehealth* OR (electronic-health* NOT electronic-health-record*) OR mhealth* OR m-health* OR telehealth* OR tele-health* OR telemedic* OR tele-medic* OR telemonitor* OR tele-monitor* OR telecare OR tele-care OR telephone* OR telepresence* OR tele-presence* OR tele-referral* OR telereferral* OR teleconsultat* OR ((digital* OR remot* OR electronic* OR virtual* OR mobile OR web-based OR webbased OR internet-based* OR internetbased*) NEAR/3 (health* OR intervention* OR consult* OR diagno* OR medicine*)) OR health-information-technolog* OR health-IoT OR h-IoT OR uhealth OR ((remot*) NEAR/3 (monitor*)) OR ((smart*) NEAR/3 (health*)) OR ((health OR medical) NEAR/2 (internet-of-things)) OR ((digital* OR remot* OR electronic* OR virtual* OR mobile) NEAR/3 (distanc*) NEAR/3 (practice*)) OR smartphone OR cellphone* OR ((smart* OR cell*) NEAR/2 (phone*)) OR videoconferenc* OR video-conferenc* OR ((video*) NEAR/3 (consult*)) OR ((mobile*) NEAR/3 (app* OR health)) OR ((home*) NEAR/3 (monitor*)) OR ((remote*) NEAR/3 (care*)) OR tele-mental* OR telemental*):ab,ti,kw) **AND** (’palliative therapy’/exp OR ’terminal care’/exp OR (palliat* OR ((symptomatic*) NEAR/3 (treat* OR therap*)) OR end-of-life* OR ((terminal*) NEAR/3 (care*))):ab,ti,kw) **AND** (’child’/exp OR ’pediatrics’/exp OR (pediatr* OR paediatr* OR neonat* OR neo-nat* OR child* OR toddler* OR boy OR boys OR girl OR girls OR infant OR infants OR baby OR babies OR newborn*):ab,ti,kw)

### Web of Science

TS=((telehospital* OR tele-hospital* OR e-health* OR ehealth* OR (electronic-health* NOT electronic-health-record*) OR mhealth* OR m-health* OR telehealth* OR tele-health* OR telemedic* OR tele-medic* OR telemonitor* OR tele-monitor* OR telecare OR tele-care OR telephone* OR telepresence* OR tele-presence* OR tele-referral* OR telereferral* OR teleconsultat* OR ((digital* OR remot* OR electronic* OR virtual* OR mobile OR web-based OR webbased OR internet-based* OR internetbased*) NEAR/2 (health* OR intervention* OR consult* OR diagno* OR medicine*)) OR health-information-technolog* OR health-IoT OR h-IoT OR uhealth OR ((remot*) NEAR/2 (monitor*)) OR ((smart*) NEAR/2 (health*)) OR ((health OR medical) NEAR/1 (internet-of-things)) OR ((digital* OR remot* OR electronic* OR virtual* OR mobile) NEAR/2 (distanc*) NEAR/2 (practice*)) OR smartphone OR cellphone* OR ((smart* OR cell*) NEAR/1 (phone*)) OR videoconferenc* OR video-conferenc* OR ((video*) NEAR/2 (consult*)) OR ((mobile*) NEAR/2 (app* OR health)) OR ((home*) NEAR/2 (monitor*)) OR ((remote*) NEAR/2 (care*)) OR tele-mental* OR telemental*) **AND** (palliat* OR ((symptomatic*) NEAR/2 (treat* OR therap*)) OR end-of-life* OR ((terminal*) NEAR/2 (care*))) **AND** (pediatr* OR paediatr* OR neonat* OR neo-nat* OR child* OR toddler* OR boy OR boys OR girl OR girls OR infant OR infants OR baby OR babies OR newborn*))

### Cochrane CENTRAL

((telehospital* OR tele NEXT/1 hospital* OR “e” NEXT/1 health* OR ehealth* OR (electronic NEXT/1 health* NOT electronic NEXT/1 health NEXT/1 record*) OR mhealth* OR “m” NEXT/1 health* OR telehealth* OR tele NEXT/1 health* OR telemedic* OR tele NEXT/1 medic* OR telemonitor* OR tele NEXT/1 monitor* OR telecare OR tele NEXT/1 care OR telephone* OR telepresence* OR tele NEXT/1 presence* OR tele NEXT/1 referral* OR telereferral* OR teleconsultat* OR ((digital* OR remot* OR electronic* OR virtual* OR mobile OR web NEXT/1 based OR webbased OR internet NEXT/1 based* OR internetbased*) NEAR/3 (health* OR intervention* OR consult* OR diagno* OR medicine*)) OR health NEXT/1 information NEXT/1 technolog* OR health NEXT/1 IoT OR “h” NEXT/1 IoT OR uhealth OR ((remot*) NEAR/3 (monitor*)) OR ((smart*) NEAR/3 (health*)) OR ((health OR medical) NEAR/2 (internet NEXT/1 of NEXT/1 things)) OR ((digital* OR remot* OR electronic* OR virtual* OR mobile) NEAR/3 (distanc*) NEAR/3 (practice*)) OR smartphone OR cellphone* OR ((smart* OR cell*) NEAR/2 (phone*)) OR videoconferenc* OR video NEXT/1 conferenc* OR ((video*) NEAR/3 (consult*)) OR ((mobile*) NEAR/3 (app* OR health)) OR ((home*) NEAR/3 (monitor*)) OR ((remote*) NEAR/3 (care*)) OR tele NEXT/1 mental* OR telemental*):ab,ti,kw) **AND** ((palliat* OR ((symptomatic*) NEAR/3 (treat* OR therap*)) OR end NEXT/1 “of” NEXT/1 life* OR ((terminal*) NEAR/3 (care*))):ab,ti,kw) **AND** ((pediatr* OR paediatr* OR neonat* OR neo NEXT/1 nat* OR child* OR toddler* OR boy OR boys OR girl OR girls OR infant OR infants OR baby OR babies OR newborn*):ab,ti,kw)

### Google Scholar

telemedicine|telehealth|telemonitoring|’tele medicine|health|consultation|monitoring’|ehealth|mhealth|’e|electronic|m|mobile|digital health’ palliation|palliative child|children

