## Appendix for "How can telehealth be used to enhance the quality of care in home-based pediatric palliative care: protocol for a systematic literature review"

20250416 Christopher Adlung

Telemedicine palliative child

SR

Trials: no

| Database searched | Platform | Years of coverage | Records | Records after duplicates removed |
|---|---|---|---|---|
| Medline ALL | Ovid | 1946 - Present | 372 | 370 |
| Embase | Embase.com | 1971 - Present | 822 | 497 |
| Web of Science Core Collection* | Web of Knowledge | 1975 - Present | 369 | 70 |
| Cochrane Central Register of Controlled Trials** | Wiley | 1992 - Present | 39 | 12 |
| Additional Search Engines: Google Scholar*** | | | 100 | 63 |
| **Total** | | | **1702** | **1012** |

*Science Citation Index Expanded (1975-present) ; Social Sciences Citation Index (1975-present) ; Arts & Humanities Citation Index (1975-present) ; Conference Proceedings Citation Index- Science (1990-present) ; Conference Proceedings Citation Index- Social Science & Humanities (1990-present) ; Emerging Sources Citation Index (2005-present)
*Exact search turned on in Web of Science Core Collection
** Manually deleted abstracts from trial registries
***Google Scholar was searched via "Publish or Perish" to download the results in EndNote.
